# Geo-social gradients in predicted COVID-19 prevalence and severity in Great Britain: results from 2,266,235 users of the COVID-19 *Symptoms Tracker app*

**DOI:** 10.1101/2020.04.23.20076521

**Authors:** Ruth C E Bowyer, Thomas Varsavsky, Carole H. Sudre, Benjamin A K Murray, Maxim B Freidin, Darioush Yarand, Sajaysurya Ganesh, Joan Capdevila, Ellen J Thompson, Elco Bakker, M. Jorge Cardoso, Richard Davies, Jonathan Wolf, Tim D Spector, Sebastien Ourselin, Claire J Steves, Cristina Menni

**Author notes:** equal contribution (Ruth C E Bowyer, Thomas Varsavsky). equal contribution (Claire J Steves, Cristina Menni).

## Abstract

Understanding the geographical distribution of COVID-19 through the general population is key to the provision of adequate healthcare services. Using self-reported data from 2,266,235 unique GB users of the *COVID Symptom Tracker app*, we find that COVID-19 prevalence and severity became rapidly distributed across the UK within a month of the WHO declaration of the pandemic, with significant evidence of “urban hot-spots”. We found a geo-social gradient associated with disease severity and prevalence suggesting resources should focus on urban areas and areas of higher deprivation. Our results demonstrate use of self-reported data to inform public health policy and resource allocation.

---

The COVID-19 epidemic has led to large-scale closures and lockdown measures across the world with the British government sanctioning lockdown from March 23^rd^ 2020.

Early in the pandemic, case distribution did not appear to be evenly spread across countries, with dense urban centres being the most affected^1^. In tandem, individuals in deprived areas have lower life expectancy^2^, are more likely to have multiple underlying comorbidities, have a higher level of influenza-associated hospitalisation^3^, and therefore could be more susceptible to COVID-19^2^. Based on the high COVID-19 prevalence in urban areas, and the known socioeconomic health gradient, we hypothesised that (i) individuals in deprived areas are at greater risk of both being exposed to and (ii) of experiencing adverse outcomes following contraction of COVID-19. Understanding the geographical distribution of the virus in a socioeconomic context is key to assist adequate healthcare resourcing, particularly intensive care beds^4^.

Here we investigated the GB geographical distribution of COVID-19 using self-reported data from over 2 million users of the *COVID-19 Symptom Tracker*^5^ app. We then identify specific geo-social, demographic and clinical factors associated with predicted COVID-19 prevalence and severity.

We studied 2,266,235 unique GB app users reporting daily on COVID-19 symptoms, hospitalisation, RT-PCR test outcomes, demographic information and pre-existing medical conditions over 24 days immediately after major social distancing measures were introduced in the GB (“lockdown”). We computed two proxies of contracting COVID-19: a predicted prevalence score^6^, (Positive Predicted Value (PPV)= 0.69[0.66; 0.71] and a predicted severity score based on symptoms associated with hospitalisation PPV=0.77(0.64;0.79).

Following aggregation of variables to local authority district level (LAD/geographic unit derived by the Office of National Statistics), we tested the geographical distribution of symptom severity and predicted prevalence using Global and Local Moran’s I tests which assess for non-random spatial distribution and clustering of a feature^7^.

We further employed linear regression adjusting for age, gender, obesity, co-morbidities and spatial autocorrelations (SAC)^8^ to assess the association between predicted COVID-19 prevalence and severity and geo-social-health factors at eight time points across 24 days using a seven-day window. Comorbidity and demographic data were included as percentage of respondents by middle super output area (MSOA, a more granular geographic unit).

The descriptive characteristics of the study population across the 8 time points are presented in **Table 1**. The number of predicted COVID-19 positive individuals ranged between 79,378 and 15,991, while the average severity (on a case 1-100) ranged from 4.16 and 1.47. Using local Moran’s I, we found that predicted COVID-19 prevalence and severity significantly clusters in urban areas across GB when considered as a proportion of the population per LAD (**Figure 1** and **Figure 1S**) adjusting for multiple testing. We also found that predicted prevalence and severity decreased over time, likely as a result of “lockdown” (**Figure 1** and **Figure 2 a** and **b**) (Pairwise Wilcoxon rank sum tests, Prevalence: all time points accept T2:T3 and T1:T4, P<0.001, Severity all time points P < 0.001). However, some hot-spots remained, suggesting in some areas social distancing measures/compliance may not have been equally effective.

**Table 1.** Demographic characteristics of the study population at 8 time points. At each time point, we only consider users who have made an assessment in the last 7 days.

|  | 29/03/2020 | 01/04/2020 | 04/04/2020 | 07/04/2020 | 10/04/2020 | 13/04/2020 | 16/04/2020 | 19/04/2020 |
| --- | --- | --- | --- | --- | --- | --- | --- | --- |
| **N** | 1,324,843 | 1,431,515 | 1,142,923 | 1083601 | 995157 | 985860 | 980608 | 1164262 |
| **Predicted COVID-19 (n/%)** | 60827<br>(4.59%) | 79378<br>(5.55%) | 62508<br>(5.47%) | 48418<br>(4.47%) | 30132<br>(3.03%) | 22352<br>(2.27%) | 16586<br>(1.69%) | 15991<br>(1.37%) |
| **Average predicted COVID-19 severity score (max 100)** | 3.72 | 4.16 | 3.77 | 3.08 | 2.38 | 2 | 1.65 | 1.47 |
| **Age, yrs** | 41.85(12.88) | 42.09(12.94) | 43.56(13.03) | 44.15(13.01) | 44.75(12.98) | 45.13(12.94) | 45.41(12.91) | 44.89(12.97) |
| **Male:** | 426,923: | 459,620: | 365,078: | 353,233: | 327,608: | 327,620: | 327,114: | 388,378: |
| **Female** | 897,920 | 971,895 | 777,845 | 730,368 | 667,549 | 658,240 | 653,494 | 775,884 |
| **Kidney Disease** | 0.60% | 0.60% | 0.60% | 0.70% | 0.70% | 0.70% | 0.70% | 0.70% |
| **Lung Disease** | 16.20% | 16.60% | 15.90% | 15.60% | 15.30% | 15.30% | 15.30% | 15.80% |
| **Diabetes** | 2.90% | 3.10% | 3.20% | 3.20% | 3.30% | 3.40% | 3.40% | 3.40% |
| **% smokers** | 13.80% | 14.00% | 12.10% | 11.50% | 10.80% | 10.60% | 10.50% | 11.10% |
| **Heart Disease** | 1.70% | 1.70% | 1.90% | 1.90% | 2.00% | 2.00% | 2.10% | 2.10% |

**Figure 1.**
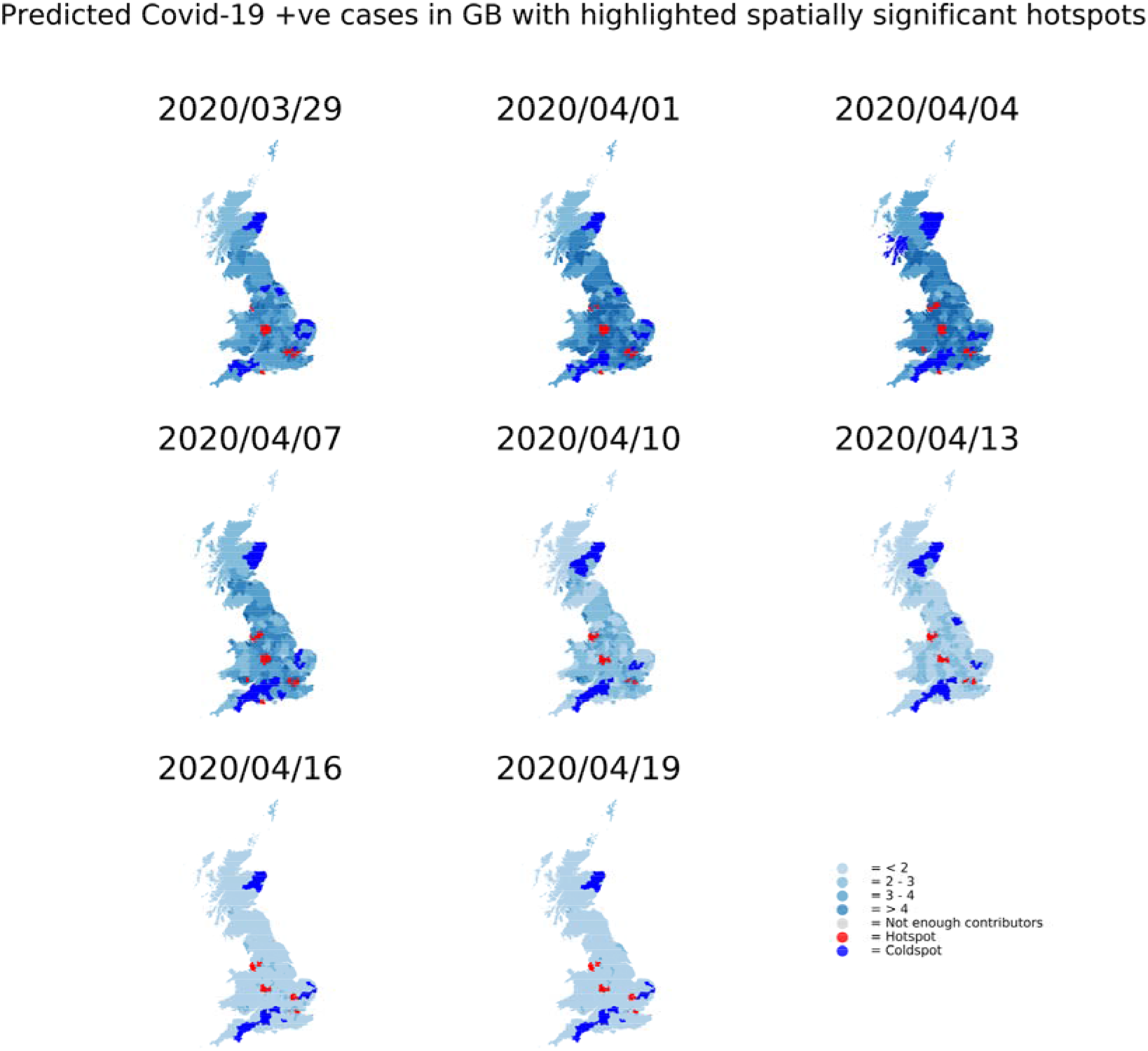
Geographical Distribution of predicted COVID-19 prevalence across eight time points. The map was created using a shapefile of Local Authority District level data from the ONS using the geopandas package and Python. Overlaid over the map are statistically significant ‘hot-spots’ and ‘cold-spots’ at LAD level. To create these regions, a ‘neighbours list was calculated from a shapefile of the LADs using a queen contiguity condition. Spatial weights were calculated assuming equal weight of neighbouring areas, and spatially lagged values which represent the average neighbouring COVID-19 prevalence/severity for each LAD. We adjusted for multiple testing using the Benjamini & Hochberg method and used the ‘spdep’ package for the spatial components of our analysis^10^. The severity score was calculated using a weighted sum of symptoms a patient had, normalised between 0 and 1. The weights were found by using the frequency of symptoms amongst a training set of 993 respondents who reported visiting hospital for a COVID-19 related issue and having a positive RT- PCR test. Predicted severity= chest pain × 0.115 + severe or significant persistent cough × 0.104 + hoarse voice × 0.097 + skipped meals × 0.096 + loss of smell × 0.092 + severe fatigue × 0.089 + shortness of breath × 0.0855 + delirium × 0.083 + fever × 0.0814 + diarrhoea × 0.081 + abdominal pain × 0.076

**Figure 2.**
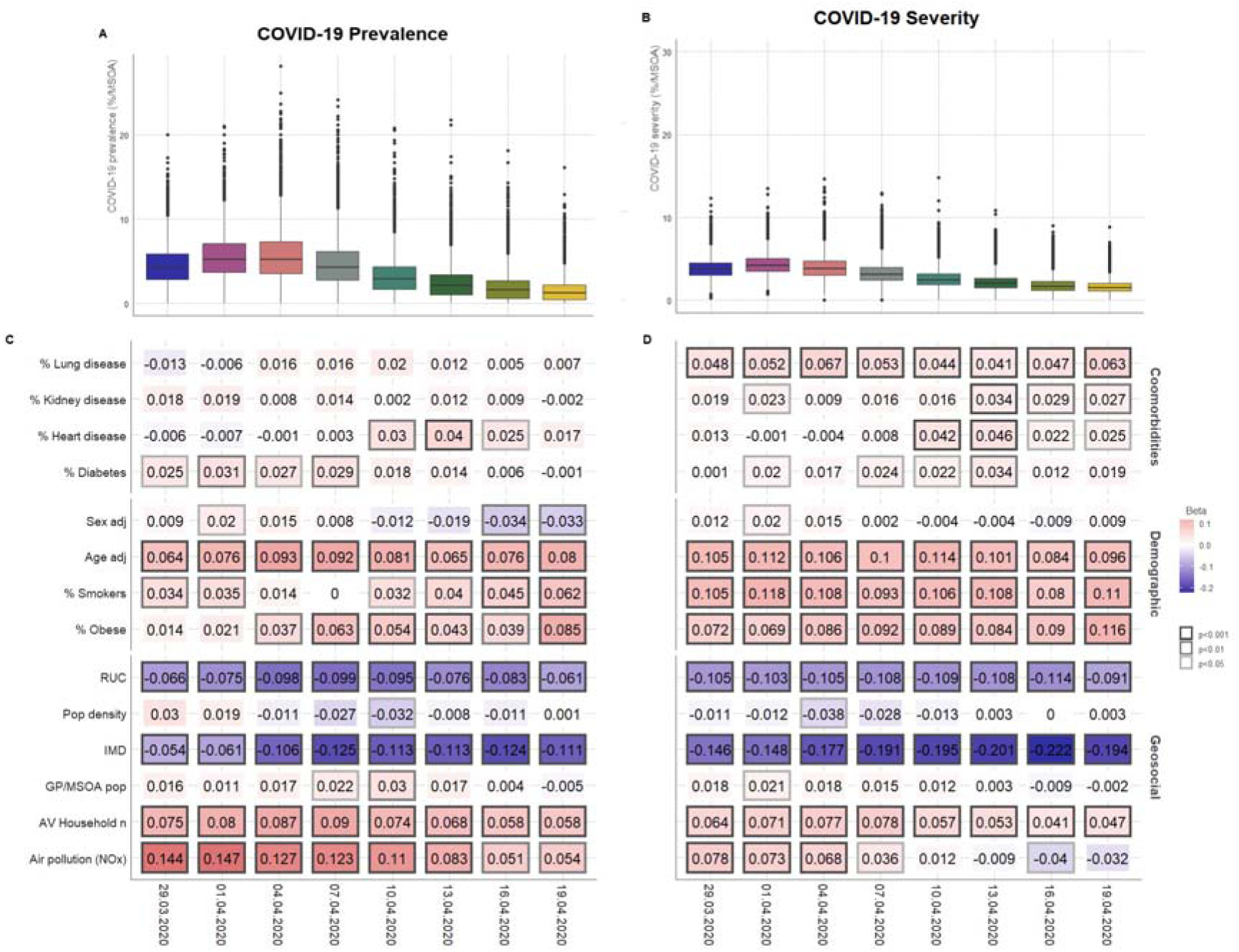
Predicted COVID- 19 A) prevalence and B) severity in the UK across eight time points. Results of multivariate linear regression models for each of the time points, where the two COVID measures C) prevalence and D) severity at MSOA level were regressed against geosocial factors: the Index of Multiple Deprivation (IMD, binned into quintiles generated on the average IMD within each MSOA, where 1 is most deprived and 5 is least), a rural-urban gradient (RUC, considered as a continuous variable where 1 is the most urban and 8 is the most rural), General practitioners per population in MSOA (GPs/MSOA, where a higher number indicates more GPs per individual by MSOA), average household number (calculated as number of inhabited dwellings/MSOA population, where a higher number indicates a higher average number of individuals per household) and comorbidity and demographic data derived from app responses representing percentage of respondents by MSOA who reported having kidney, heart or lung disease, and who are diabetic, a smoker or obese (calculated as BMI<30). Finally, we derived age and sex variables to adjust for response bias and considered as demographic factors: these were calculated as the difference of the expected mean/ratio of age/sex in the MSOA (derived from ONS population data) and the observed mean/ratio of age/sex amongst respondents. Therefore, a positive number suggests we had *less* males/*younger* respondents than would be expected were our population to be the same as the average population by MSOA, and thus a positive association with COVID prevalence/severity suggests greater association with this reporting bias. A lagged variable was included to account for spatial autocorrelation. Only MSOAs where at least 20 individuals were considered (n = 8097). Standardised coefficients are reported. Analysis conducted in RStudio v1.1.423 and R v3.6.3.

In the more granular analysis, we find that urbanicity, area-level deprivation and average household size were positively associated to higher predicted COVID-19 prevalence and severity (*P*<0.01) across all time points; with stronger associations for severity than for prevalence (**Figure 2 c** and **d**). This suggests that people in deprived and/or urban areas remained at higher risk of more severe symptoms. Moreover, we see a positive trend between NOx pollution and COVID-19 prevalence and to a lesser extent severity.

Finally, as expected, we find the association of obesity, smoking and lung disease to be stronger with predicted COVID-19 severity than prevalence.

Here, we observe that predicted COVID-19 prevalence and severity is significantly higher in urban areas compared to rural, and in more deprived areas compared to less deprived. This could reflect the likelihood of individuals in more deprived areas to work in vocations, or live with people who work in vocations, where they are unable to work from home and are thus more likely to be exposed to circulating COVID-19. Accumulation of socio-environmental exposures across the life course are known to contribute to a greater health deficit and disease burden^2^; our results suggest that COVID- 19 is no exception.

Moreover, our study illustrates how app data could be used to successfully monitor COVID-19 over time and identify hotspots as the viral pandemic progresses and social distancing measures are implemented or eased. Using this method, we detected a geo-social gradient associated with disease severity and prevalence in the context of COVID-19 suggesting resources should focus on urban areas, areas with highest deprivation, higher average household number and higher air pollution.

Our study has some limitations and assumptions. First, we used self-reported data. Second, volunteers using the app are a self-selected group that may not be fully representative of the general population. Third, our data on COVID-19 incidence is not from confirmed tests via RT-PCR, but rather from two variables predicted based on user responses. Additionally, we assume that people who suffer from COVID-19 are equally likely to use the app as those who do not. We also assume that people will report the symptoms in the same way. Finally, we aggregated data at MSOA level as we did not have enough respondents for more granular geography.

Future work could seek to integrate this data with data on area-level morbidity, extended pollution data and ethnicity. Indeed higher mortality has been observed amongst minority ethnic groups^9^ and disentangling the environmental and biological factors contributing to greater disease burden in both deprived areas and among ethnic minorities is an essential focus of future work to ensure resources and intervention are better assigned.

## Data Availability

Anonymised research data will be shared with third parties via the centre for Health Data Research UK (HDRUK.ac.uk). Data updates can be found on https://covid.joinzoe.com

## Conceived and designed the experiments

CJS, TDS, SO, CM;

## Analysed the data

RCEB, TV.

## Contributed reagents/materials/analysis tools

MF, CHS, BM, MBF, DY, SG, JC, EJT,EB, MJC, RD, JW

## Wrote the manuscript

RCEB, TV, CM

## Revised the manuscript

all

## Acknowledgements

This work was supported by Zoe Global. The Department of Twin Research is funded by the Wellcome Trust, Medical Research Council, European Union, Chronic Disease Research Foundation (CDRF), Zoe Global Ltd and the National Institute for Health Research (NIHR)-funded BioResource, Clinical Research Facility and Biomedical Research Centre based at Guy’s and St Thomas’ NHS Foundation Trust in partnership with King’s College London. CM is funded by the Chronic Disease Research Foundation and by the MRC Aim-Hy project grant. CHS is an Alzheimer’s Society Junior Fellowship AS-JF-17-011; SO and MJC are funded by the Wellcome/EPSRC Centre for Medical Engineering (WT203148/Z/16/Z), Wellcome Flagship Programme (WT213038/Z/18/Z).

We express our sincere thanks to all the participants of the COVID Symptom Tracker app. We thank the staff of Zoe Global Limited, the Department of Twin Research for their tireless work in contributing to the running of the study and data collection.

## Ethics

The Ethics for the app has been approved by KCL ethics Committee and all users provided consent for non-commercial use. An informal consultation with TwinsUK members over email and social media prior to the app having been launched found that they were overwhelmingly supportive of the project.

## Figure Legends

**Figure S1.**
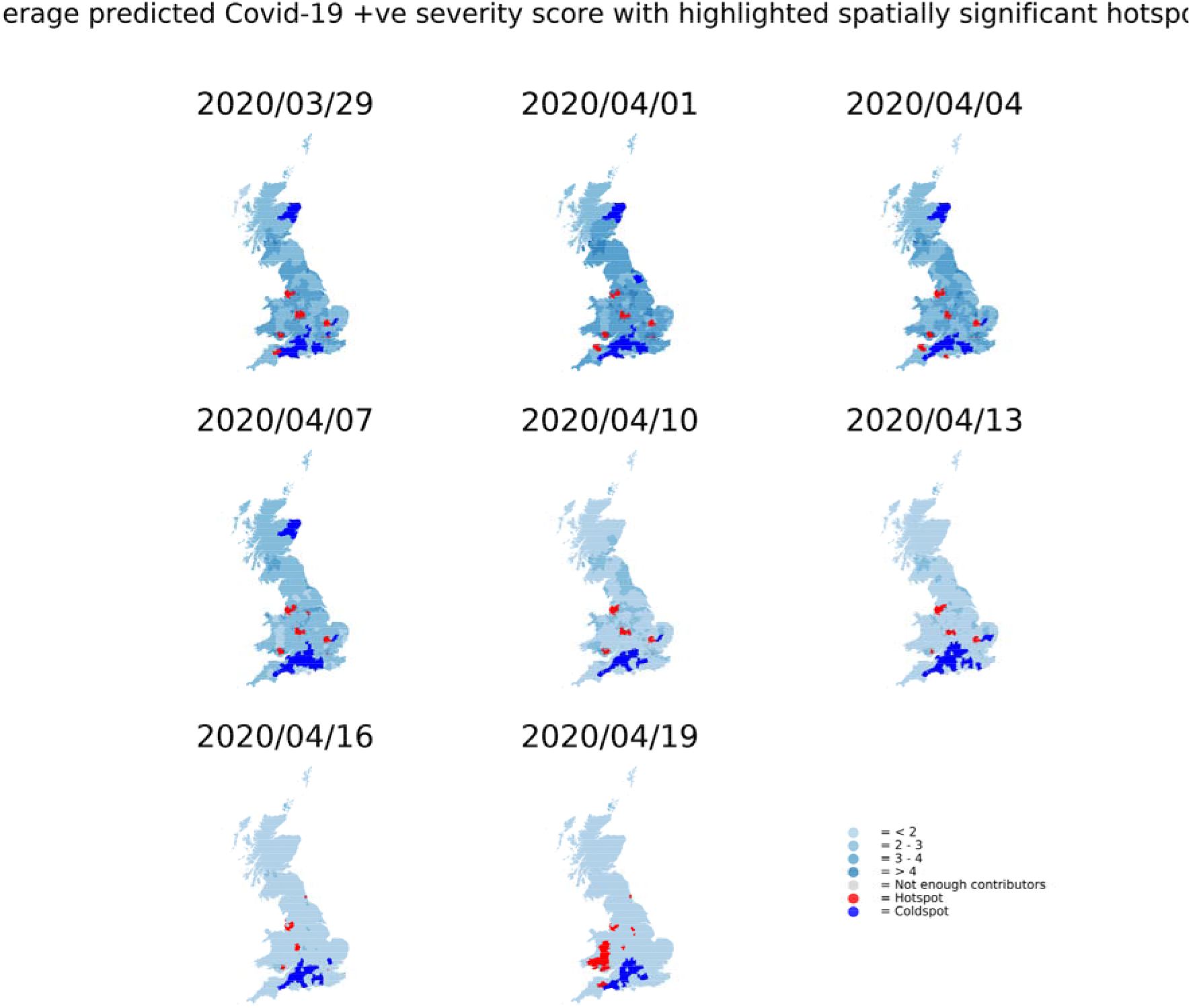
Geographical Distribution of predicted COVID-19 severity across eight time points.

